# A room, a bar and a classroom: how the coronavirus is spread through the air depends on heavily mask filtration efficiency

**DOI:** 10.1101/2020.11.10.20227710

**Authors:** Devabhaktuni Srikrishna

## Abstract

**Background:** Recently the US CDC acknowledged by that the COVID-19 crisis is facilitated at least in part by aerosolized virus exhaled by symptomatic, asymptomatic, or pre-symptomatic infected individuals. Disposable N95 masks remain in short supply due to their use in healthcare settings during the Coronavirus pandemic, whereas NIOSH-approved elastomeric N95 (eN95) masks remain immediately available for use by essential workers and the general public. New reusable N95 mask options with symmetric filtration efficiency can be anticipated to be NIOSH approved in the coming months, today’s eN95 masks have asymmetric filtration efficiency upon inhalation (95%) and exhalation (well under 95%) but are available now during the Fall and Winter when Coronavirus cases are expected to peak.

**Methods:** Based on the Wells-Riley model of infection risk, we examine how the rate of transmission of the virus from one infected person in a closed, congested room with poor ventilation to several other susceptible individuals is impacted by the filtration efficiency of the masks they are wearing. Three scenarios are modeled – a room (6 people, 12’ × 20’ × 10’), a bar (18 people, 20’ × 40’ × 10’), and a classroom (26 people, 20’ × 30’ × 10’) with one infectious individual and remaining susceptibles. By dynamically estimating the accumulation of virus in aerosols exhaled by the infected person in these congested spaces for four hours using a “box model,” we compare the transmission risk (probability) when susceptible people based on a realistic hypothesis of face mask protection during inhaling and exhaling e.g. using cloth masks or N95 respirators.

**Results:** Across all three scenarios, cloth masks modeled with 30% symmetric filtration efficiency alone were insufficient to stop the spread (18% to 40% infection risk), whereas eN95 masks (modeled as 95% filtration efficiency on inhalation, 30% on exhalation) reduced the infection risk to 1.5% to 3.6%. Symmetric filtration of 80% reduces the risk to 1.7% to 4.1% and symmetric filtration of 95% would further reduce the risk to 0.11% to 0.26%.

**Conclusion:** This modeling of mask filtration efficiency suggests that the pandemic could be readily controlled within several weeks if (in conjunction with sensible hygiene) a sufficiently large majority of people wear asymmetric but higher-filtration masks (e.g. eN95) that also block aerosols whenever exposed to anyone else outside their household in order to completely stop inter-household spread.

## Introduction

Recently a newspaper El Pais illustrated the aerosolized spread of Coronavirus in closed, congested environments with poor ventilation (a room, a bar, and a classroom) based on the estimator that models aerosol spread of Coronavirus. They concluded that cloth masks and increasing ventilation together could help reduce the spread [10]. Whereas the effect of using masks with better filtration efficiency is a personal choice that can provide uniform protection wherever the wearer happens to go, but the effect mask filtration efficiency has on transmission risk not been modeled in these closed, congested, poorly ventilated scenarios.

In this report, we model what would happen if a single intervention of masks with asymmetric and symmetric filtration were to be used. We find that with asymmetric N95 masks the transmission risk could be brought near zero. In addition, blocking both inhalation and exhalation at 95% virtually eliminated any infections from aerosols (e.g. with a disposable or symmetric N95). For full details of the aerosol transmission model, estimation, and input parameters please see the Methods section.

## Results

First we verified that transmission indeed remains significant with cloth masks (modeled as blocking 30% on both inhalation and exhalation of aerosols) while ventilation remains poor. Then instead of cloth masks, we modeled all participants using N95 masks that block 95% of inhaled aerosols and only 30% of exhaled aerosols using an elastomeric N95 with an exhale valve (e.g. valve covered with a cloth mask). In all scenarios (room, bar, classroom), the infected person (labeled as “patient 0” in the illustrations below) is modeled as exhaling 60 infectious doses of virus per hour and the participants are present for four hours (240 minutes). All other values in the model were set to their defaults from the “classroom” scenario in the estimator.

### Room (1^st^ scenario)

As shown below, six people are present in a 12 foot by 20 foot room with poor ventilation (0.5 air changes per hour). With no masks three more people are infected. Using cloth masks two more people are expected to be infected, whereas with asymmetric N95 masks no one else is expected to be infected (3.6% chance of infection). Symmetric filtration above 80% reduces risk similar to asymmetric N95.

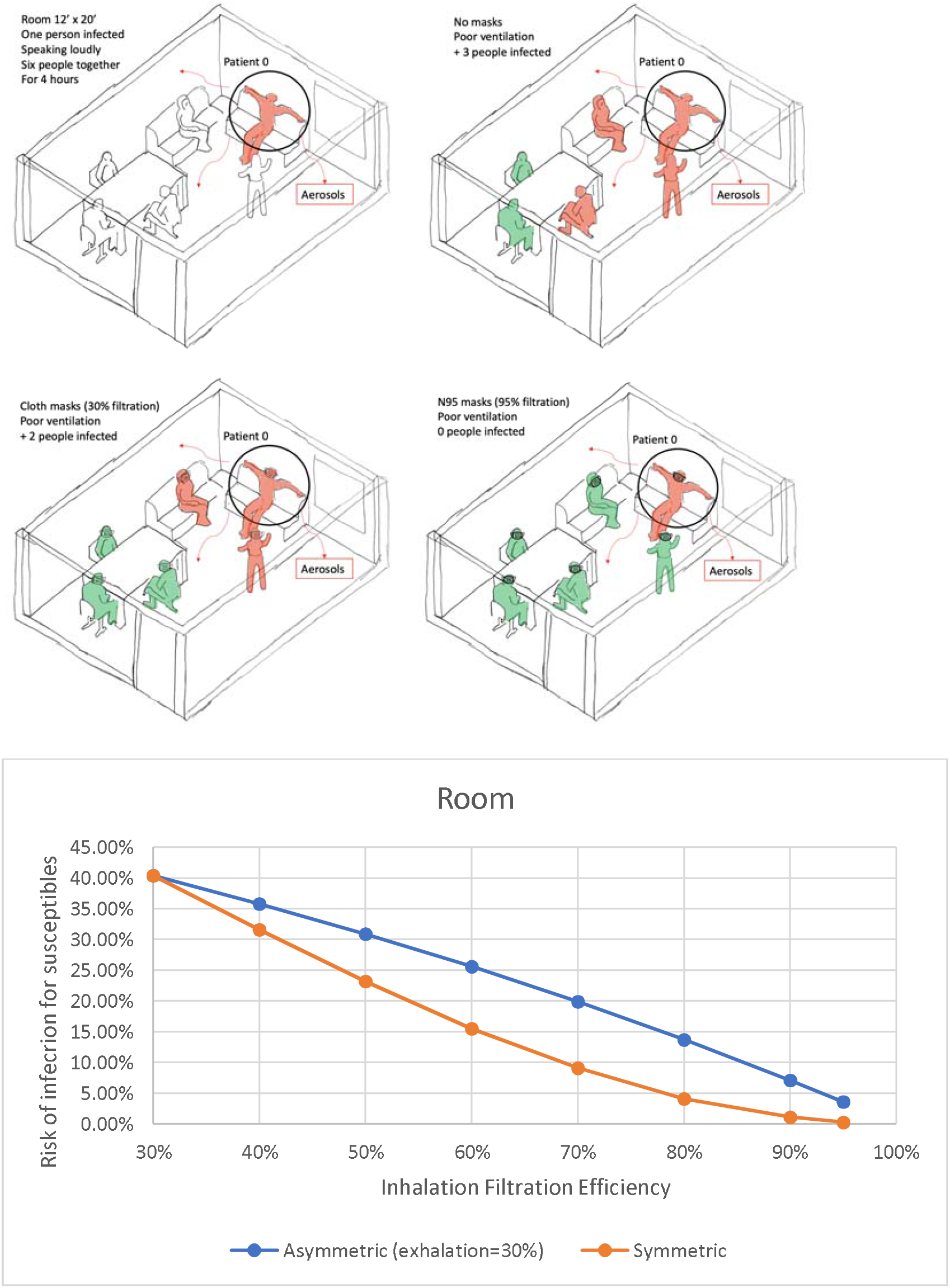

### Bar (2^nd^ scenario)

As shown below, 18 people are present in a 20 foot by 40 foot bar with no ventilation and windows shut (0 air changes per hour). With no masks five more people are infected. Using cloth masks three more people are expected to be infected, whereas with asymmetric N95 masks no one else is expected to be infected (1.5% chance of infection). Symmetric filtration above 80% reduces risk similar to asymmetric N95.

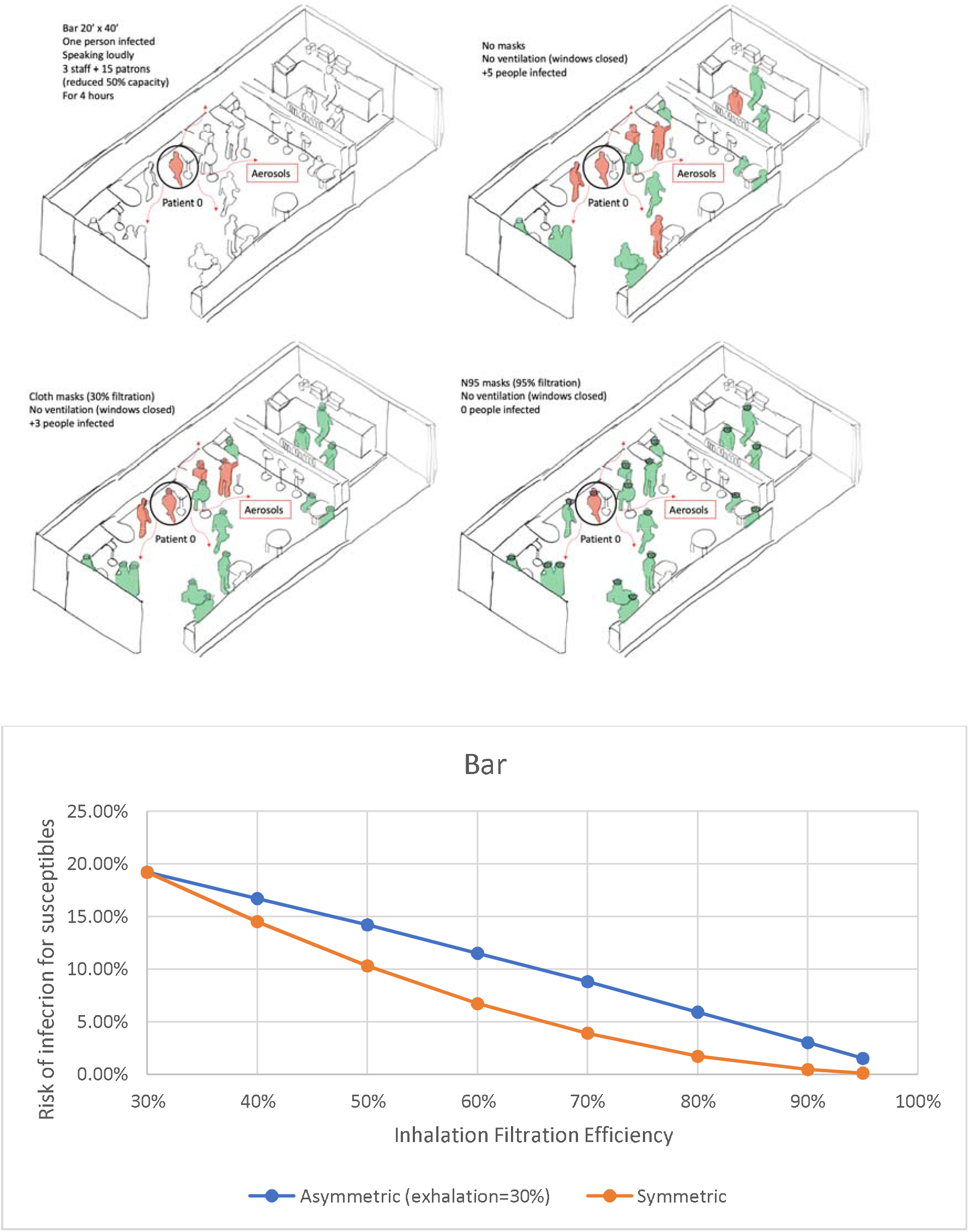

### Classroom (3^rd^ scenario)

As shown below, 25 students and one teacher (who is infected) are present in a 20 foot by 30 foot classroom with poor ventilation (0.5 air changes per hour). With no masks nine students end up infected. Using cloth masks five more people are expected to be infected, whereas with asymmetric N95 masks no one else is expected to be infected (1.5% chance of infection). Symmetric filtration above 80% reduces risk similar to asymmetric N95.

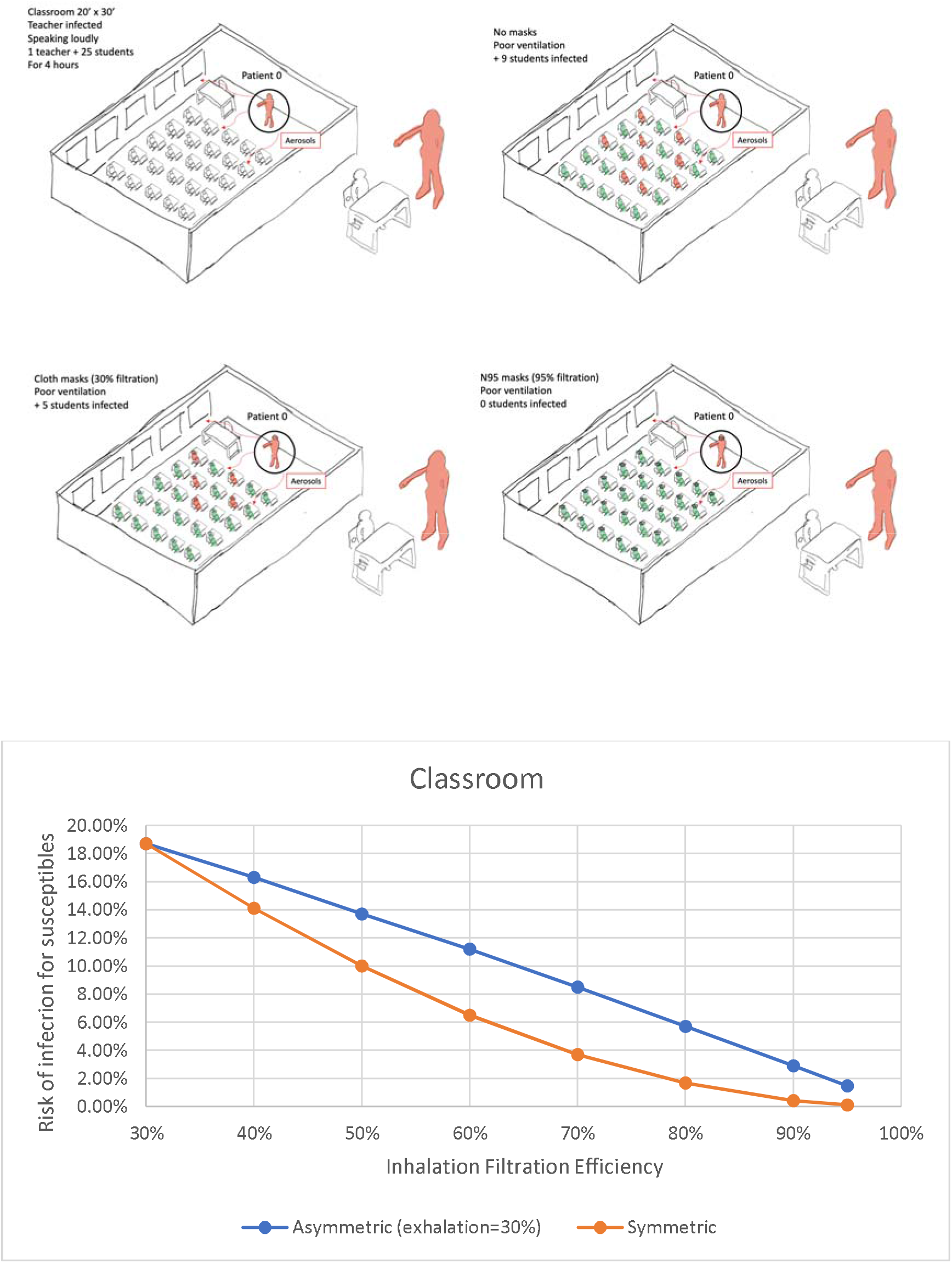

## Discussion

This modeling suggests the pandemic would probably be over within several weeks if (in addition to sensible hygiene) a sufficiently large majority of people were to wear high-filtration masks that also block aerosols whenever exposed to anyone else outside their household (such as elastomeric N95 with cloth mask covering the valve), especially in closed, congested environments that lack good ventilation.

The US CDC has stated that Coronavirus is spread mainly by respiratory droplets [1], and they leave as an open question how often it is transmitted by aerosol versus other means such as fomite or droplet [2]. In [3], Lee et al. make three fundamental observations:

1. Although intra-household spread is numerically higher (often target of immense contact tracing resources), the pandemic cannot be sustained without inter-household spread.
2. Intra-household contacts are so long and intense that it is immaterial whether droplets, fomites (contaminated surfaces), or aerosolized particles are primarily responsible because all have ample opportunity.
3. Inter-household there are greater variety of infectious contacts in public, at work, at school, in social environments where aerosolized super-spreading is likely to be more prevalent.

Preventing intra-household spread can be intractable in many cases due to the close nature of the inter-personal interactions, whereas using social protection [4] it may be much more feasible to eliminate inter-household spread without the economic burden of lockdowns. These non-pharmaceutical interventions [5] most likely include better masks that block aerosols and sensible hygiene which in combination need to be sufficiently protective to prevent seeding of new household infections so that any pre-existing intra-household transmission eventually runs its course after several cycles.

Aerosols are not effectively blocked by most cloth masks that are widely used today [6]. Even surgical masks issued to passengers on a large ship did not prevent 59% of them from being infected [7]. Cloth masks and surgical masks may mitigate spread or reduce symptoms [8] but they may not stop transmission especially in closed, congested environments with poor ventilation. Although N95 masks are known to block aerosols with 95% filtration efficiency (or equivalently reduction of the aerosols by a factor of 20x), public health agencies continue to discourage N95 use [12] due to a persistent narrative of shortage [11] of disposable N95 masks used by healthcare workers. These disposable N95 masks are now available online [13], but if not worn with a proper fit [14], the level of protection can decline making them less effective [15]. Compared to a disposable N95 that can leak, a “stretchy” elastomeric N95 (eN95) is more likely to fit better [16], is not supply-constrained [17] in the near-term, approved by CDC/NIOSH for N95-level protection [18], and manufacturable at scale and readily available for $20 to $80 each [19].

During Coronavirus crisis, the CDC abruptly ruled out [21] use of any masks designed with unidirectional (exhale) valves [22] as being insufficiently protective [23] to others nearby which includes many industrial N95 masks such as eN95. The rationale is if the person wearing the mask happens to be infected with Coronavirus the air breathed out [24] of a unidirectional valve is unfiltered [25] and could infect others unless everyone else nearby is wearing one too (by comparison aerosols also flow out of typical cloth masks). Newer models expected in 2021 or beyond aim to resolve this by incorporating bidirectional N95 filtering of both inhaled and exhaled air [26] [27]. While waiting for such innovations to gain regulatory approval from the federal government (CDC-NIOSH), workarounds to comply with local laws [28] for currently available eN95 models include supplementary face covering as demonstrated by San Francisco’s health officer [29], sealing the valve using tape [30], or a surgical mask over an eN95 as proposed by the CDC [31].

Unlike disposable N95, many healthcare institutions remain reluctant to use eN95 [39] so they are not in short supply and available online. However the eN95 are more efficient, reusable, and scalable. In a recent study comparing eN95 to disposable N95, the cost was, conservatively, 10x less per month and after a month no healthcare workers wanted to return to disposables [20]. Numerous manufacturers include Envomask ([32] and first image below), MSA, Honeywell, and for illustration, a 3M elastomeric mask [33] with P95 filter ([34] and second image below) costs $23 on Amazon (P100 [35] offers more protection with less breathability). With 10 replacement filters that are each rated for 40 hours of continuous use and could potentially be reused (rotated) for significantly longer [36], that comes to under $100 per person or under $40 billion to equip the US population.

Although there is no near-term shortage of eN95, scaling up manufacturing of N95 filters to the population will eventually require investment in expensive machinery which is currently unattractive for investors to finance since demand for such masks typically dries up after an epidemic is over [37]. Stable, growing, long-term consumer demand for N95s can attract private investment in new manufacturing lines just as it does for smartphones [38]. Whereas by telling consumers not to use N95s public health leaders may unwittingly exacerbate shortages by undermining private investment in N95 manufacturing [12].

Covid-19 is not the worst virus that could confront humanity. There are ∼200 known species of human RNA viruses, and on average, two or more new ones are discovered annually [40] [41]. High-quality, daily-use respirators (masks) that filter out aerosols were originally conceptualized 25 years ago to prepare soldiers for biological warfare [42]. If daily-use respirators are available on-hand for everyone to put on whenever there is knowledge of bioaerosols in the air, it would instantly slow down if not stop the spread of any virus or bacteria transmitted by aerosols. Today such respirators are readily available to consumers in the form of elastomeric N95 (eN95) [43], and perhaps nothing else can provide as much individual protection to a wide range of bioaerosol threats.

## Methods

Based on the Wells-Riley infection model, we examine how the rate of transmission of the virus from one infected person in a closed, congested room with poor ventilation to several other susceptible individuals is impacted by the filtration efficiency of the masks they are wearing. By dynamically estimating the accumulation of virus in aerosols exhaled by the infected person in these congested spaces using a “box model” we compare the transmission risk (probability) when susceptible people uses cloth masks or N95 respirators based on a realistic hypothesis of face mask protection during inhaling and exhaling.

Based on the development in [44], the average number of quanta (infectious doses of virus) per unit volume ‘C_avg_’ in a closed, congested room with poor ventilation over a duration ‘D’ can be modeled as the average of the time-dependent concentration of the virus in the room’s air ‘C(t)’. C(t) in turn depends on the emission rate ‘E’ of quanta per unit time from the infected person, the volume of the room ‘V’, and combined rate of loss of infectious virus from the air ‘L’ (per unit time) due to the effect of ventilation, decay of the virus viability over time, or loss of viruses onto surfaces. This leads to a first order differential equation with terms for increase E/V and decrease L C(t):

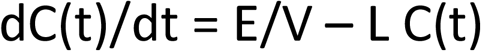

The solution to this differential equation is

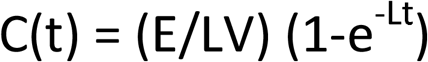

The average of C(t) over the duration of time D is

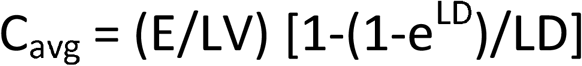

The average number of quanta inhaled ‘n’ by a susceptible person can be estimated based on C_avg_, the breathing rate of the susceptible people ‘B’, duration D, inhalation filtration efficiency of the mask worn by the susceptible ‘x’, and exhalation filtration efficiency of the mask worn by the infected person ‘y’:

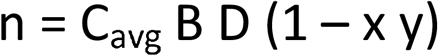

We use the above equations to estimate the probability (risk) P as a function of mask filtration efficiency. The Wells-Riley model uses the Poisson distribution to stochastically estimate the risk ‘P’ (probability) of a susceptible person being infected by inhaling an infectious dose (quanta) of the virus that is present in the aerosols exhaled by one infected person where the average number of quanta inhaled would be ‘n’ in a given period of time:

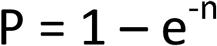

We modeled three poorly ventilated scenarios (room with 6 people, bar with 18 people, classroom with 25 people) each with one infected person who is speaking loudly based on the estimation methodology in Jimenez et. al [9].

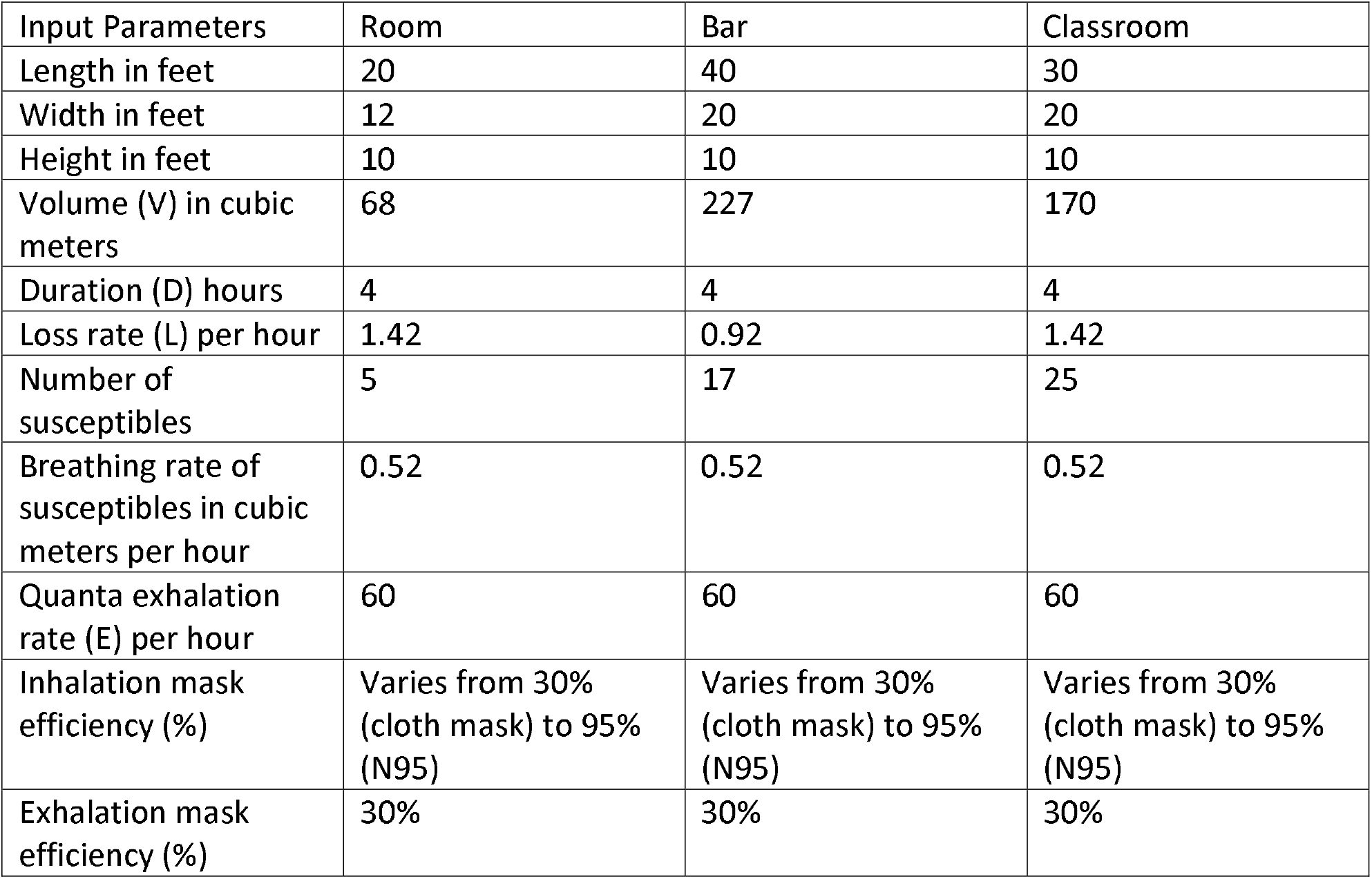

## Data Availability

All data used is available in the manuscript

## References

[1] https://www.cdc.gov/coronavirus/2019-ncov/prevent-getting-sick/how-covid-spreads.html

[2] https://www.cdc.gov/coronavirus/2019-ncov/more/scientific-brief-sars-cov-2.html

[3] https://science.sciencemag.org/content/370/6515/406

[4] https://www.medrxiv.org/content/10.1101/2020.04.01.20049981v3

[5] https://www.barrons.com/articles/masks-are-critical-in-fighting-covid-19-they-need-federal-research-dollars-51603456511

[6] https://advances.sciencemag.org/content/6/36/eabd3083

[7] https://thorax.bmj.com/content/75/8/693

[8] https://www.nejm.org/doi/full/10.1056/NEJMp2026913

[9] http://tinyurl.com/covid-estimator

[10] https://english.elpais.com/society/2020-10-28/a-room-a-bar-and-a-class-how-the-coronavirus-is-spread-through-the-air.html

[11] https://www.washingtonpost.com/graphics/2020/local/news/n-95-shortage-covid/

[12] https://www.fda.gov/medical-devices/personal-protective-equipment-infection-control/n95-respirators-surgical-masks-and-face-masks

[13] https://www.sfgate.com/shopping/article/guide-to-purchasing-KN95-and-NIOSH-approved-masks-15550519.php

[14] https://aip.scitation.org/doi/figure/10.1063/5.0022968#v4

[15] https://www.statnews.com/2020/04/16/n95-masks-training-needed-protect-against-covid-19/

[16] https://safety.blr.com/workplace-safety-news/employee-safety/respiratory-protection/CalOSHAissues-updated-ATD-respirator-guidance/

[17] https://www.federalregister.gov/documents/2020/09/14/2020-20115/a-national-elastomeric-half-mask-respirator-ehmr-strategy-for-use-in-healthcare-settings-during-an

[18] https://www.cdc.gov/coronavirus/2019-ncov/hcp/elastomeric-respirators-strategy/index.html

[19] https://hbr.org/2020/10/essential-workers-need-better-masks

[20] https://www.journalacs.org/article/S1072-7515(20)30471-3/fulltext

[21] https://www.cdc.gov/coronavirus/2019-ncov/prevent-getting-sick/cloth-face-cover-guidance.html#masks-with-vents

[22] https://www.washingtonpost.com/health/2020/08/13/cdc-mask-guidance-masks-valves/

[23] https://www.ajicjournal.org/article/S0196-6553(20)30815-4/fulltext

[24] https://www.tandfonline.com/doi/full/10.1080/02786826.2020.1812502

[25] https://www.ncbi.nlm.nih.gov/pmc/articles/PMC7521346/

[26] https://www.maskforce1.com/

[27] https://openstandardrespirator.org/

[28] https://sf.gov/information/masks-and-face-coverings-coronavirus-pandemic

[29] https://taragonmd.github.io/2020/08/22/respirator-and-face-covering-to-protect-against-coronavirus-and-poor-air-quality/

[30] https://www.sfgate.com/science/article/Cloth-masks-won-t-protect-you-from-wildfire-15496106.php

[31] https://blogs.cdc.gov/niosh-science-blog/2020/09/08/elastomeric/

[32] https://envomask.com/

[33] https://www.amazon.com/3M-Thermoplastic-Elastomer-Respirator-Connection/dp/B00W632ITA

[34] https://www.amazon.com/3M-Particulate-Filter-2071-filters/dp/B0013ZCVLW/

[35] https://www.amazon.com/3M-50051131070009-Particulate-Filter-2091/dp/B07571LKP4/

[36] https://www.cdc.gov/niosh/topics/hcwcontrols/recommendedguidanceextuse.html

[37] https://www.washingtonpost.com/business/scarcity-of-raw-material-still-squeezes-n95-mask-makers/2020/09/10/94586834-f31e-11ea-8025-5d3489768ac8_story.html

[38] https://www.washingtonpost.com/outlook/2020/05/21/why-dont-hospitals-have-enough-masks-because-coronavirus-broke-market/

[39] https://www.mountsinai.org/files/MSHealth/Assets/HS/About/Coronavirus/MSHS_ProperUseF aceCovering-June24-2020.pdf

[40] https://www.ncbi.nlm.nih.gov/pmc/articles/PMC6157708/

[41] https://www.nytimes.com/video/health/100000007293397/covid-pandemics-causes-documentary.html

[42] https://pubmed.ncbi.nlm.nih.gov/8855053/

[43] https://www.csfa.net/CSFA/CalFF/articles/Coronavirus_and_wildfire_particles_are_in_the_air__Elastomeric_N95__eN95__can_help.aspx

[44] https://www.medrxiv.org/content/10.1101/2020.06.15.20132027v2

